# Internalized HIV stigma, antiretroviral therapy adherence, and health-related quality of life among women living with HIV in Nigeria

**DOI:** 10.64898/2026.09.28.26364230

**Authors:** Dolapo O. Raji, Folahanmi T. Akinsolu, Lilian O. Ezechi, Olunike R. Abodunrin, Titilola A. Gbajabiamila, Tomilola Musari-Martin, Abdulrasheed Oba, Lateef A. Blessing, Inumidun A. Olatunji, Gift M. Adalaja, Justin N. Brai, Posi E. Aduroja, Afolakemi O. Oredein, Zanthia Wiley, Oliver C. Ezechi

## Abstract

**Background:** Internalized HIV stigma remains a major psychosocial challenge for women living with HIV (WLHIV) and may adversely affect antiretroviral therapy (ART) adherence and health-related quality of life (HRQoL). However, fewer studies in sub-Saharan Africa have examined how internalized stigma and ART adherence jointly relate to multidimensional HRQoL among WLHIV receiving routine HIV care. This study examined the associations of internalized HIV stigma and ART adherence with HRQoL among WLHIV in Lagos, Nigeria, and explored whether the relationship between stigma and HRQoL varied by ART adherence status.

**Methods:** This was a baseline analytical study nested within a quasi-experimental study among WLHIV receiving care at the HIV Treatment Centre of the Nigerian Institute of Medical Research, Lagos, Nigeria. The analysis included 1,005 women with complete baseline data on internalized stigma, ART adherence, HRQoL, and selected covariates. Internalized stigma was assessed using the 10-item Internalized Stigma of HIV/AIDS Tool and categorized as low or high using the sample median. ART adherence was measured using pharmacy refill adherence and self-reported adherence, which were combined into a composite overall adherence variable. HRQoL was assessed using the WHOQOL-HIV BREF, and the primary outcome was the total domain-based quality-of-life score derived from the six WHOQOL-HIV domains. Linear regression was used to examine associations between stigma and adherence and the total domain-based QoL score, and an interaction term between internalized stigma and overall ART adherence was included to assess effect modification.

**Results:** The mean age of participants was 47.8 years (SD 12.5), and 56.0% had high levels of internalized stigma. Overall, 73.3% were classified as adherent using the composite ART adherence measure. HRQoL was consistently poorer among women with high internalized stigma across all WHOQOL-HIV BREF domains. Women with high internalized stigma had a substantially lower total domain-based QoL score than those with low stigma (106.5; SD 11.0 vs 117.8; SD 10.6; p < 0.001). In the final multivariable model, high internalized stigma remained the strongest independent correlate of lower QoL (β = -10.07, 95% CI: -11.41 to -8.73; p < 0.001), while overall ART non-adherence was also associated with lower QoL (β = -2.78, 95% CI: -4.24 to -1.33; p < 0.001). Older age and living more than 5 km from the health facility were associated with lower QoL, whereas social support was associated with higher QoL. Interaction analyses showed that predicted QoL was highest among women with low stigma who were adherent and lowest among those with high stigma who were non-adherent.

**Conclusions:** Internalized HIV stigma and ART non-adherence were independently associated with poorer HRQoL among WLHIV in Lagos, Nigeria. Internalized stigma emerged as the strongest correlate of poorer well-being, underscoring the need to move beyond narrowly biomedical models of HIV care. Interventions that integrate stigma reduction, adherence support, and psychosocial strengthening may be particularly valuable for improving patient-centered outcomes among WLHIV in routine HIV care settings.

## Introduction

The scale-up of antiretroviral therapy (ART) has transformed HIV infection from a life-threatening disease into a chronic, manageable condition.^1–4^ As survival has improved, the goals of HIV care have expanded beyond viral suppression and mortality reduction to include broader patient-centered outcomes such as health-related quality of life (HRQoL).^5–8^ This shift is particularly important for women living with HIV (WLHIV), whose experiences of illness and care are shaped not only by clinical factors but also by gendered social expectations, caregiving responsibilities, disclosure pressures, and persistent stigma.^9,10^ Globally, 40.8 million people were living with HIV (PLHIV) in 2024, and women and girls accounted for 53% of all PLHIV.^11,12^ In Nigeria, women continue to bear a disproportionate burden of the epidemic, with national data showing substantially higher HIV prevalence among women than men of reproductive age (UNAIDS, 2025; NACA, 2020).^1,11,13,14^

Among the psychosocial factors influencing HIV outcomes in women, internalized stigma remains one of the most consequential.^15,16^ Internalized HIV stigma occurs when WLHIV absorb negative societal attitudes and direct them inward, resulting in shame, guilt, reduced self-worth, social withdrawal, and fear of disclosure.^17^ For many women, these experiences are layered onto existing social and gender-related vulnerabilities, including dependency within relationships, fear of abandonment, caregiving burden, and concern about social judgment.^18–20^ As a result, internalized stigma may undermine psychological well-being, weaken social relationships, and disrupt sustained engagement in care.^16,21,22^

HRQoL has become an increasingly important outcome in HIV care because it captures how individuals function physically, psychologically, socially, and spiritually while living with a chronic condition.^23,24^ The WHOQOL-HIV BREF was specifically developed to assess multidimensional well-being among PLHIV.^23,24^ Previous studies have shown that stigma is negatively associated with multiple QoL domains, and that this burden may be particularly pronounced among WLHIV.^15,25^ For example, among WLHIV in India, stigma was linked to lower HRQoL and poorer treatment-related functioning,^26,27^ while studies from Europe have similarly shown that self-stigma is negatively associated with HRQoL across several domains.^25^ These findings suggest that stigma is not only a social experience for WLHIV, but also an important determinant of day-to-day wellbeing.^25–27^

ART adherence may also influence HRQoL directly, through better health stability and symptom control, and indirectly, through greater confidence in treatment and continuity of care.^23,24^ However, stigma and adherence are unlikely to operate independently. Among WLHIV, internalized stigma may reduce QoL directly while also influencing treatment behavior through fear, secrecy, emotional distress, and disengagement from support.^15,26^ This is especially relevant in Nigeria, where stigma remains a persistent barrier to testing, treatment uptake, adherence, and broader psychosocial well-being despite the expansion of HIV services.^22^

Although stigma, adherence, and QoL have each been studied separately, fewer studies have examined how internalized stigma and ART adherence jointly relate to multidimensional HRQoL among WLHIV in sub-Saharan African care settings.^15,26,28^ This gap is important because WLHIV may experience HIV-related stigma in distinct ways that shape both treatment behavior and lived well-being.^15,26^ A clearer understanding of these interrelationships may help inform more integrated psychosocial and adherence-support strategies within routine HIV care, particularly in low-resource settings where women often navigate layered clinical, social, and economic vulnerabilities.^28,29^

This study examined the associations of internalized HIV stigma and ART adherence with HRQoL among WLHIV receiving care in Lagos, Nigeria. It also explored whether the relationship between internalized stigma and HRQoL varied according to ART adherence status. By focusing on women receiving routine HIV care in a sub-Saharan African setting, this study provides context-specific evidence to inform more integrated psychosocial and adherence-support interventions.

## Methods

### Study design

This study was a baseline analytical study nested within a quasi-experimental study among WLHIV receiving care in Lagos, Nigeria. For the present analysis, only data collected during the pre-intervention phase were used to examine the associations of internalized HIV stigma and ART adherence with HRQoL. Restricting the analysis to baseline data enabled assessment of these relationships prior to exposure to the intervention.^30–32^

### Study setting

The study was conducted at the HIV Treatment Centre of the Nigerian Institute of Medical Research (NIMR), Lagos, Nigeria, a high-volume facility providing comprehensive HIV care, including clinical management, laboratory monitoring, adherence support, and follow-up services.^33,34^ The centre was selected because it serves a large and diverse population of WLHIV and offers the routine care infrastructure necessary for participant recruitment, clinical record abstraction, and collection of psychosocial and treatment-related data.^35,36^

### Study population

The study population comprised women living with HIV aged 18 years and above who were receiving care at the HIV Treatment Centre, NIMR, Lagos, Nigeria.^35,36^ Participants were receiving routine HIV care at the study site and constituted the source population for the baseline assessment.

### Study eligibility

Women were eligible for inclusion if they were aged 18 years or older, currently enrolled for HIV care at the NIMR HIV Treatment Centre, had been on ART for at least 6 months, were able to provide written informed consent, and were willing to participate in the baseline assessment. Women were excluded if they were younger than 18 years, were not receiving care at the study clinic, had severe cognitive impairment or untreated psychiatric illness that could affect their ability to participate meaningfully, were concurrently enrolled in another intervention study targeting stigma, adherence, or quality of life, or declined consent.

### Study recruitment

Participants were recruited consecutively during routine clinic visits by trained research assistants. After eligibility screening, the study objectives and procedures were explained, and written informed consent was obtained from women who agreed to participate. Baseline data were collected through interviewer-administered questionnaires in a private setting. The questionnaire captured socio-demographic information, clinical and treatment history, internalized stigma, ART adherence, and quality of life, while selected clinical parameters were abstracted from routine records where available. The present analysis included 1,005 women with complete baseline data on internalized stigma, ART adherence, quality of life, and selected covariates.

### Data collection procedures

Data were collected at baseline using a structured interviewer-administered questionnaire and abstraction of selected clinical information from routine medical records. The questionnaire was organized into socio-demographic, clinical, stigma, adherence, and quality-of-life sections. Interviews were conducted face-to-face in a private setting by trained research assistants to ensure confidentiality and data completeness. Clinical information, including viral load, CD4 count, WHO clinical stage, ART regimen, and anthropometric measures, was abstracted from participants’ records where available. (See Supplementary File 1)

#### a) Socio-demographic and clinical data

Socio-demographic information collected included age, marital status, education, employment status, monthly income, religion, ethnic group, number of children, living arrangement, residential area, distance to the nearest health facility, mode of transportation to the clinic, health insurance status, disclosure of HIV status, and source of health-related support. Clinical and treatment-related information included time since HIV diagnosis, current ART use, duration on ART, recent viral load category, missed doses in the past 30 days, hospitalization for HIV-related illness in the preceding 6 months, ART side effects, clinic appointment attendance, receipt of adherence or psychosocial support, current CD4 count, most recent viral load, WHO clinical stage, current ART regimen, and history of opportunistic infection in the previous 6 months. Anthropometric data included height and weight, from which body mass index (BMI) was calculated as weight in kilograms divided by height in metres squared.^37,38^ BMI was categorized as underweight (<18.5 kg/m²), normal weight (18.5–24.9 kg/m²), overweight (25.0–29.9 kg/m²), and obese (≥30.0 kg/m²).^38–40^

#### b) Internalized HIV stigma

Internalized stigma was assessed using the Internalized Stigma of HIV/AIDS Tool (ISAT), a 10-item measure of negative self-perceptions related to living with HIV.^17^ The scale captures feelings such as shame, guilt, worthlessness, concealment, inferiority, self-blame, and perceived burden. In this study, each item was rated on a 5-point Likert scale ranging from 1 (“strongly disagree”) to 5 (“strongly agree”), with higher scores indicating greater internalized stigma. Item scores were summed to obtain a total stigma score. For descriptive and regression analyses, stigma scores were dichotomized at the sample median into low internalized stigma (<20) and high internalized stigma (≥20).^17,41^

#### c) ART adherence

ART adherence was assessed using two complementary approaches: pharmacy refill adherence and self-reported adherence.^42,43^

- Pharmacy refill adherence was derived from pharmacy refill records and reflected the proportion of prescribed ART refills obtained within the expected refill period. A continuous pharmacy refill adherence score was first computed and converted to a percentage.^44^ For descriptive purposes, refill adherence was categorized as good (score ≥2.5; approximately ≥83%), fair (score 1.5–2.0; approximately 50%–67%), and poor (score <1.5; <50%). For binary analyses, pharmacy refill adherence was dichotomized as adherent (≥95%) or non-adherent (<95%).^44–46^
- Self-reported adherence was assessed using structured questions on missed doses, medication-taking consistency, and timing of doses.^42,47^ Responses were scored on a 0–4 scale. For descriptive purposes, scores ≥3.5 were classified as adherent and scores <3.5 as non-adherent. For the primary binary adherence measure, participants were classified as adherent if they reported never missing a dose, or missing no more than one dose in the previous month, taking all or most of their medication as prescribed, and taking doses on time or within one hour of the prescribed time. Participants who did not meet all these criteria were classified as non-adherent. A composite overall ART adherence variable was then generated, with participants classified as adherent only if they met the ≥95% threshold on both pharmacy refill and self-reported adherence measures. This combined approach was used to strengthen adherence ascertainment and reduce reliance on a single measurement method.^42^

#### d) Quality of life

HRQoL was assessed using the WHOQOL-HIV BREF, a 31-item instrument developed for use among people living with HIV and structured across six domains: physical health, psychological health, level of independence, social relationships, environment, and spirituality/religion/personal beliefs.^23,24^ The instrument also includes two general items assessing overall QoL and general health. Responses were recorded on Likert-type scales, and negatively phrased items were reverse-coded in accordance with standard scoring procedures. Domain scores were calculated by averaging the constituent item scores within each domain and multiplying the mean by four, yielding domain scores ranging from 4 to 20, with higher scores indicating better QoL.^23,24^ For this analysis, the primary outcome was the total domain-based QoL score, calculated as the sum of the six domain scores, while the two general items were summarized descriptively and not included in multivariable modelling.

#### e) Covariates

Potential covariates were selected a priori based on their conceptual relevance to internalized stigma, ART adherence, and health-related quality of life, as well as evidence from prior literature on determinants of HIV-related wellbeing.^24,48,49^ These included age, marital status, education level, monthly income, religion, number of children, living arrangement, residential area, health facility distance, health insurance status, disclosure status, support status, time since HIV diagnosis, current ART use, recent viral load category, most recent viral load, WHO clinical stage, opportunistic infection history, and BMI category. Disclosure status was categorized as disclosed versus not disclosed. Support status was categorized as support versus no support. Residential area was categorized as rural, semi-urban, or urban, and health facility distance was categorized as less than 1 km, 1–5 km, or more than 5 km.

### Statistical analysis

All statistical analyses were conducted using R version 4.4.1. Continuous variables were summarized using means and standard deviations, while categorical variables were summarized using frequencies and percentages. Participant characteristics were compared across internalized stigma categories using chi-squared tests for categorical variables and two-sample t-tests for continuous variables.

Bivariate associations among internalized stigma scores, adherence variables, and quality-of-life measures were examined using Spearman’s rank correlation coefficients, given the ordinal nature of several measures and the potential for non-normal distributions.^50^

The primary inferential analysis examined the associations of internalized HIV stigma and overall ART adherence with total domain-based quality-of-life score using linear regression.^51^ Candidate covariates were identified a priori based on conceptual relevance to stigma, adherence, and quality of life.^48^ In Model 1, each candidate predictor was entered separately to estimate unadjusted associations with total domain-based QoL score. Variables associated with the outcome at p ≤ 0.05 in unadjusted analyses were entered simultaneously into Model 2 to estimate adjusted associations. Variables that remained statistically significant at p ≤ 0.05 in Model 2 were retained in Model 3, which served as the final parsimonious model. Effect estimates were reported as beta coefficients (β) with 95% confidence intervals (CIs), and statistical significance was defined as a two-sided p-value <0.05.

To assess whether the association between internalized HIV stigma and total domain-based QoL score differed according to ART adherence status, an interaction term between stigma category (low vs high) and overall ART adherence (adherent vs non-adherent) was introduced into a multivariable linear regression model.^51^ The interaction model was specified as:

**QoL = β₀ + β₁(stigma category) + β₂(ART adherence) + β₃(stigma category × ART adherence) + ε**

where β₃ represents the interaction effect between internalized HIV stigma and ART adherence on total domain-based QoL score. Adjusted predicted means and 95% confidence intervals were then estimated for each stigma–adherence combination and presented graphically to aid interpretation of the interaction. Additional details on variable definitions, scoring procedures, and model-building strategy are provided in Supplementary File 2.

### Ethical considerations

Ethical approval for the study was obtained from the Nigerian Institute of Medical Research Institutional Review Board (IRB-25-030) before participant recruitment and data collection. Written informed consent was obtained from all participants before enrolment. Participation was voluntary, and participants were informed that refusal to participate or withdrawal at any time would not affect their access to HIV care or treatment services.

Because the study involved women living with HIV, special attention was given to privacy and confidentiality. Interviews were conducted in a private setting by trained research assistants to reduce the risk of inadvertent HIV status disclosure and to ensure participants could respond freely to potentially sensitive questions on stigma, disclosure, adherence, and quality of life. Participants were also informed that they could decline to answer any question they found uncomfortable.

To protect confidentiality, questionnaires and clinical data were linked using study identification codes rather than personal identifiers. All data were de-identified prior to analysis, and access to study data was restricted to authorized members of the research team. Overall, the study was conducted in a manner that respected participants’ dignity, autonomy, privacy, and wellbeing throughout the research process.

## Results

### Participant characteristics

A total of 1,005 women living with HIV (WLHIV) were included in the analysis. The mean age was 47.8 years (SD 12.5). Overall, 66.3% were married, 49.5% had secondary education, and 37.4% had tertiary education. Most participants resided in urban areas (71.8%), 54.1% lived within 1–5 km of the health facility, 96.1% had disclosed their HIV status, 79.3% reported having support, and 97.0% were currently receiving ART. More than half of the participants (56.0%) were classified as having high internalized stigma. Compared with WLHIV with low internalized stigma, those with high internalized stigma differed significantly by monthly income, residential area, distance to the health facility, disclosure status, social support, latest viral load result, and overall ART adherence. Detailed baseline characteristics are presented in Table 1. Expanded baseline characteristics are presented in Supplementary Table S1.

**Table 1:** Participant characteristics by internalized stigma category among WLHIV.

| Characteristic | High internalized stigma (n = 564)<br>N (%) | Low internalized stigma (n = 441)<br>N (%) | Overall (N = 1,005)<br>N (%) | p-value |
| --- | --- | --- | --- | --- |
| **Age, years** | 47.3 (12.3) | 48.4 (12.7) | 47.8 (12.5) | 0.159<sup>b</sup> |
| **Marital status** |  |  |  | 0.264<sup>a</sup> |
| Married | 365 (64.7) | 301 (68.3) | 666 (66.3) |  |
| Separated/divorced | 35 (6.2) | 17 (3.9) | 52 (5.2) |  |
| Single | 93 (16.5) | 76 (17.2) | 169 (16.8) |  |
| Widowed | 71 (12.6) | 47 (10.7) | 118 (11.7) |  |
| **Educational level** |  |  |  | 0.217<sup>a</sup> |
| No formal education | 19 (3.4) | 9 (2.0) | 28 (2.8) |  |
| Primary education | 66 (11.7) | 38 (8.6) | 104 (10.3) |  |
| Secondary education | 271 (48.0) | 226 (51.2) | 497 (49.5) |  |
| Tertiary education | 208 (36.9) | 168 (38.1) | 376 (37.4) |  |
| **Monthly income** |  |  |  | <0.001<sup>a**</sup> |
| Less than ₦10,000 | 33 (5.9) | 54 (12.2) | 87 (8.7) |  |
| ₦10,000–₦49,999 | 87 (15.4) | 70 (15.9) | 157 (15.6) |  |
| ₦50,000–₦99,999 | 115 (20.4) | 117 (26.5) | 232 (23.1) |  |
| ₦100,000 and above | 99 (17.6) | 104 (23.6) | 203 (20.2) |  |
| Prefer not to say | 230 (40.8) | 96 (21.8) | 326 (32.4) |  |
| **Residential area** |  |  |  | <0.001<sup>a**</sup> |
| Rural | 58 (10.3) | 11 (2.5) | 69 (6.9) |  |
| Semi-urban | 108 (19.1) | 106 (24.0) | 214 (21.3) |  |
| Urban | 398 (70.6) | 324 (73.5) | 722 (71.8) |  |
| **Distance to health facility** |  |  |  | 0.003<sup>a**</sup> |
| <1 km | 135 (23.9) | 143 (32.4) | 278 (27.7) |  |
| 1–5 km | 312 (55.3) | 232 (52.6) | 544 (54.1) |  |
| >5 km | 117 (20.7) | 66 (15.0) | 183 (18.2) |  |
| **BMI category** |  |  |  | 0.326<sup>a</sup> |
| Underweight | 24 (4.3) | 17 (3.9) | 41 (4.1) |  |
| Normal weight | 228 (40.4) | 158 (35.8) | 386 (38.4) |  |
| Overweight | 173 (30.7) | 158 (35.8) | 331 (32.9) |  |
| Obese | 139 (24.6) | 108 (24.5) | 247 (24.6) |  |
| **HIV status disclosed** |  |  |  | 0.012<sup>a**</sup> |
| Yes | 534 (94.7) | 432 (98.0) | 966 (96.1) |  |
| No | 30 (5.3) | 9 (2.0) | 39 (3.9) |  |
| **Social support** |  |  |  | <0.001<sup>a**</sup> |
| No support | 144 (25.5) | 64 (14.5) | 208 (20.7) |  |
| Any support | 420 (74.5) | 377 (85.5) | 797 (79.3) |  |
| **Time since HIV diagnosis** |  |  |  | 0.646<sup>a</sup> |
| <6 months | 10 (1.8) | 4 (0.9) | 14 (1.4) |  |
| 6–12 months | 5 (0.9) | 5 (1.1) | 10 (1.0) |  |
| 13–24 months | 8 (1.4) | 5 (1.1) | 13 (1.3) |  |
| >24 months | 541 (95.9) | 427 (96.8) | 968 (96.3) |  |
| **Currently on ART** |  |  |  | 0.105<sup>a</sup> |
| No | 12 (2.1) | 18 (4.1) | 30 (3.0) |  |
| Yes | 552 (97.9) | 423 (95.9) | 975 (97.0) |  |
| **Latest viral load result** |  |  |  | 0.019<sup>a**</sup> |
| Undetectable (<50 copies/mL) | 316 (56.0) | 285 (64.6) | 601 (59.8) |  |
| Suppressed (50–999 copies/mL) | 30 (5.3) | 27 (6.1) | 57 (5.7) |  |
| Not suppressed (≥1,000 copies/mL) | 11 (2.0) | 9 (2.0) | 20 (2.0) |  |
| Don't know | 207 (36.7) | 119 (27.0) | 326 (32.4) |  |
| **Overall ART adherence** |  |  |  | 0.043<sup>a**</sup> |
| Adherent | 399 (70.7) | 338 (76.6) | 737 (73.3) |  |
| Non-adherent | 165 (29.3) | 103 (23.4) | 268 (26.7) |  |

<sup>a</sup> Chi-squared test; <sup>b</sup> Two-sample t-test; <sup>c</sup> ANOVA; <sup>\*\*</sup> Significant p-values (≤0.05)

### Distribution of internalized stigma and ART adherence

Figure 1 shows the distribution of internalized stigma and ART adherence measures in the study population. Among the 1,005 WLHIV included in the analysis, 56.0% had high internalized stigma and 44.0% had low internalized stigma. Adherence estimates varied by method of assessment: pharmacy refill records classified 87.8% of participants as adherent, whereas self-reported adherence classified 73.8% as adherent. Using the composite measure requiring concordance between pharmacy refill and self-reported adherence, 73.3% of participants were classified as adherent and 26.7% as non-adherent.

**Figure 1:**
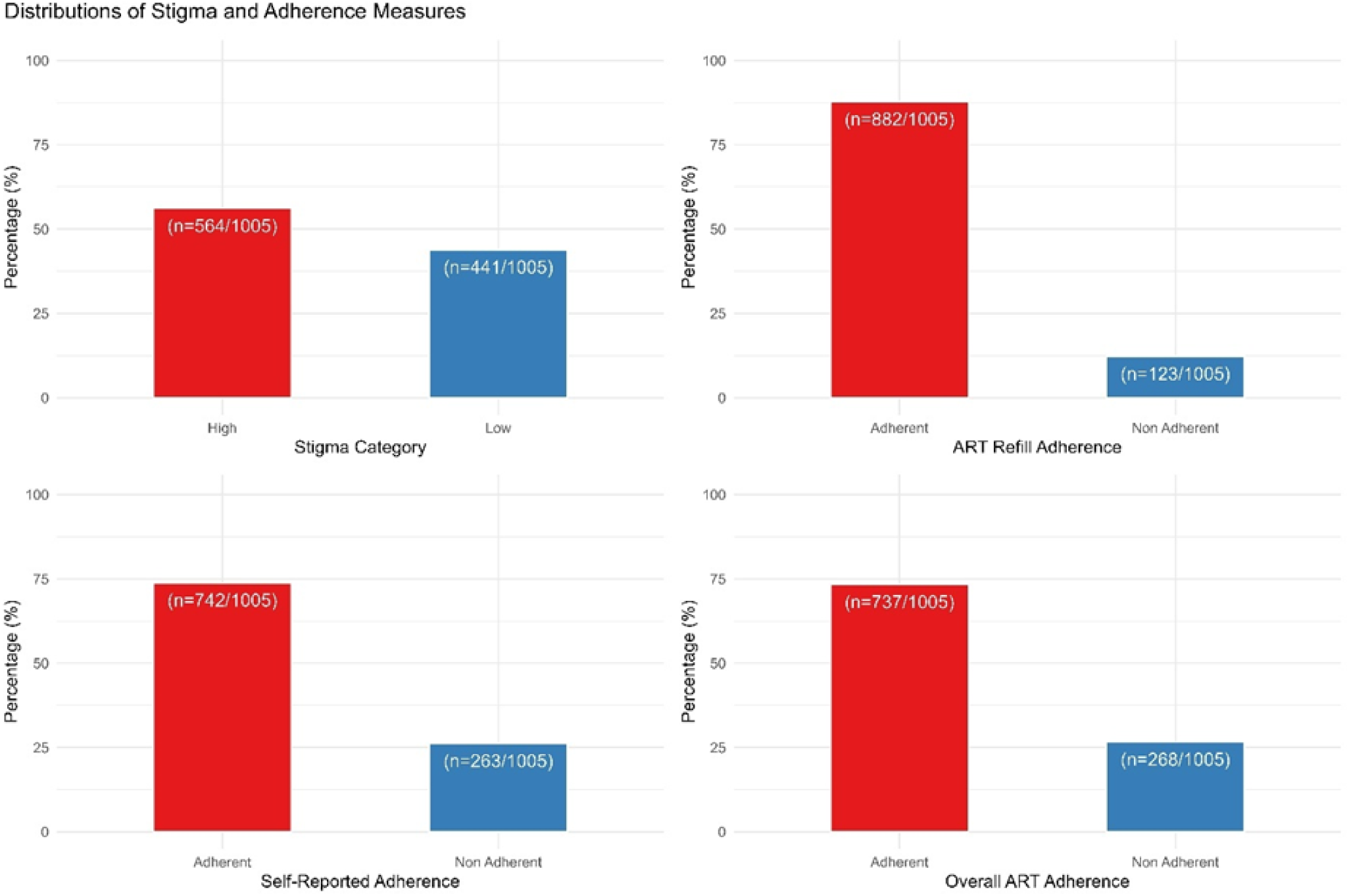
Distribution of internalized stigma and ART adherence measures among women living with HIV

### Quality of life by internalized stigma status

Table 2 summarizes domain-specific and total domain-based quality-of-life scores according to internalized stigma category. Health-related quality of life was consistently poorer among WLHIV with high internalized stigma than among those with low internalized stigma. Across the WHOQOL-HIV BREF domains, women with high stigma had significantly lower mean scores for physical health, psychological health, social relationships, level of independence, environment, and spirituality/religion/personal beliefs (all p < 0.001). The largest absolute differences were observed in the psychological, environmental, and spirituality/religion/personal beliefs domains. Importantly, WLHIV with high internalized stigma had a substantially lower total domain-based QoL score than those with low stigma (106.5, SD 11.0 vs 117.8, SD 10.6, p < 0.001).

**Table 2.** WHOQOL-HIV BREF domain scores and total domain-based quality-of-life score by internalized stigma category.

| Outcome | High internalized stigma (n = 564) | Low internalized stigma (n = 441) | Overall (N = 1,005) | p-value |
| --- | --- | --- | --- | --- |
| Physical health | 15.7 (2.3) | 17.8 (2.1) | 16.6 (2.5) | <0.001 |
| Psychological health | 15.1 (2.2) | 17.6 (2.0) | 16.2 (2.5) | <0.001 |
| Social relationships | 16.2 (1.7) | 17.8 (1.9) | 16.9 (2.0) | <0.001 |
| Level of independence | 14.9 (2.2) | 15.6 (2.7) | 15.2 (2.4) | <0.001 |
| Environment | 14.6 (2.2) | 16.4 (2.4) | 15.4 (2.5) | <0.001 |
| Spirituality/religion/personal beliefs | 14.5 (3.3) | 17.5 (2.9) | 15.8 (3.4) | <0.001 |
| **Total domain-based QoL score** | **106.5 (11.0)** | **117.8 (10.6)** | **111.4 (12.2)** | **<0.001** |

**Table 3.** Multivariable linear regression of factors associated with total domain-based quality-of-life score among women living with HIV.

| Predictor | Category / comparison | Adjusted $\beta$ (95% CI) | p-value |
| --- | --- | --- | --- |
| Age | Per 1-year increase | -0.10 (-0.15 to -0.04) | <0.001** |
| BMI category | Underweight (Ref) |  |  |
|  | Normal weight vs underweight | 3.62 (1.29 to 5.94) | 0.002** |
|  | Overweight vs underweight | -2.13 (-4.00 to -0.27) | 0.025** |
|  | Obese vs underweight | 1.26 (-0.01 to 2.53) | 0.052 |
| Distance to health facility | 1–5 km (Ref) |  |  |
|  | <1 km vs 1–5 km | 1.97 (0.47 to 3.48) | 0.010** |
|  | >5 km vs 1–5 km | -4.66 (-6.43 to -2.90) | 0.001** |
| Monthly income | ~~N~~10,000–~~N~~49,999 (Ref) |  |  |
|  | ~~N~~100,000 and above vs Ref | 1.28 (-0.88 to 3.43) | 0.245 |
|  | ~~N~~50,000–~~N~~99,999 vs Ref | -2.45 (-4.54 to -0.36) | 0.021** |
|  | Less than ~~N~~10,000 vs Ref | -2.26 (-5.09 to 0.58) | 0.118 |
|  | Prefer not to say vs Ref | -2.22 (-4.22 to -0.21) | 0.030** |
| Opportunistic infection history | Yes vs No | 4.72 (1.97 to 7.47) | <0.001** |
| Social support | Any support vs no support | 2.27 (0.65 to 3.89) | 0.006** |
| Overall ART adherence | Non-adherent vs adherent | -2.78 (-4.24 to -1.33) | <0.001** |
| Internalized stigma | High vs low | -10.07 (-11.41 to -8.73) | <0.001** |

\*\* indicates statistical significance at $p < 0.05$ .
$\beta$ coefficients represent adjusted mean differences in total domain-based QoL score. Positive coefficients indicate higher QoL scores, whereas negative coefficients indicate lower QoL scores.

### Factors associated with total domain-based quality of life

In unadjusted linear regression analyses, high internalized stigma and overall ART non-adherence were both associated with lower total domain-based QoL score. Other variables associated with QoL at the unadjusted level included age, monthly income, distance to the health facility, residential area, BMI category, number of children, opportunistic infection history, and social support. Unadjusted and intermediate regression models are presented in Supplementary Table S2.

In the final multivariable model, internalized stigma remained the strongest independent correlate of poorer quality of life. Compared with WLHIV with low internalized stigma, those with high internalized stigma had a 10.07-point lower total domain-based QoL score (β = -10.07, 95% CI: -11.41 to -8.73; p < 0.001). Overall ART non-adherence was also independently associated with poorer quality of life, with non-adherent women having a 2.78-point lower total domain-based QoL score than adherent women (β = -2.78, 95% CI: -4.24 to -1.33; p < 0.001).

Several additional covariates were independently associated with total domain-based QoL score. Older age was associated with a modest reduction in QoL (β = -0.10, 95% CI: -0.15 to -0.04; p < 0.001). Relative to the reference BMI category, normal weight was associated with higher QoL (β = 3.62, 95% CI: 1.29 to 5.94; p = 0.002), whereas overweight was associated with lower QoL (β = -2.13, 95% CI: -4.00 to -0.27; p = 0.025). Compared with living 1–5 km from the health facility, living less than 1 km away was associated with higher QoL (β = 1.97, 95% CI: 0.47 to 3.48; p = 0.010), while living more than 5 km away was associated with lower QoL (β = -4.66, 95% CI: -6.43 to -2.90; p = 0.001). WLHIV earning ₦50,000–₦99,999 and those who preferred not to disclose their income had lower QoL scores than the reference income group (β = -2.45, 95% CI: -4.54 to -0.36; p = 0.021; and β = -2.22, 95% CI: -4.22 to -0.21; p = 0.030, respectively). Social support was positively associated with QoL (β = 2.27, 95% CI: 0.65 to 3.89; p = 0.006). A history of opportunistic infection was also positively associated with total domain-based QoL score in the adjusted model (β = 4.72, 95% CI: 1.97 to 7.47; p < 0.001).

### Interaction between internalized stigma and ART adherence on total domain-based quality of life

Figure 2 presents the adjusted predicted total domain-based QoL scores by internalized stigma category and overall ART adherence. Predicted QoL was highest among WLHIV with low internalized stigma who were adherent to ART and lowest among those with high internalized stigma who were non-adherent. Within both adherence strata, WLHIV with high internalized stigma had lower predicted QoL than those with low internalized stigma. Similarly, within both stigma strata, adherent women had higher predicted QoL than non-adherent women. The greater separation between the low- and high-stigma groups among adherent WLHIV suggests that the association between internalized stigma and total domain-based QoL scores may vary by adherence status.

**Figure 2.**
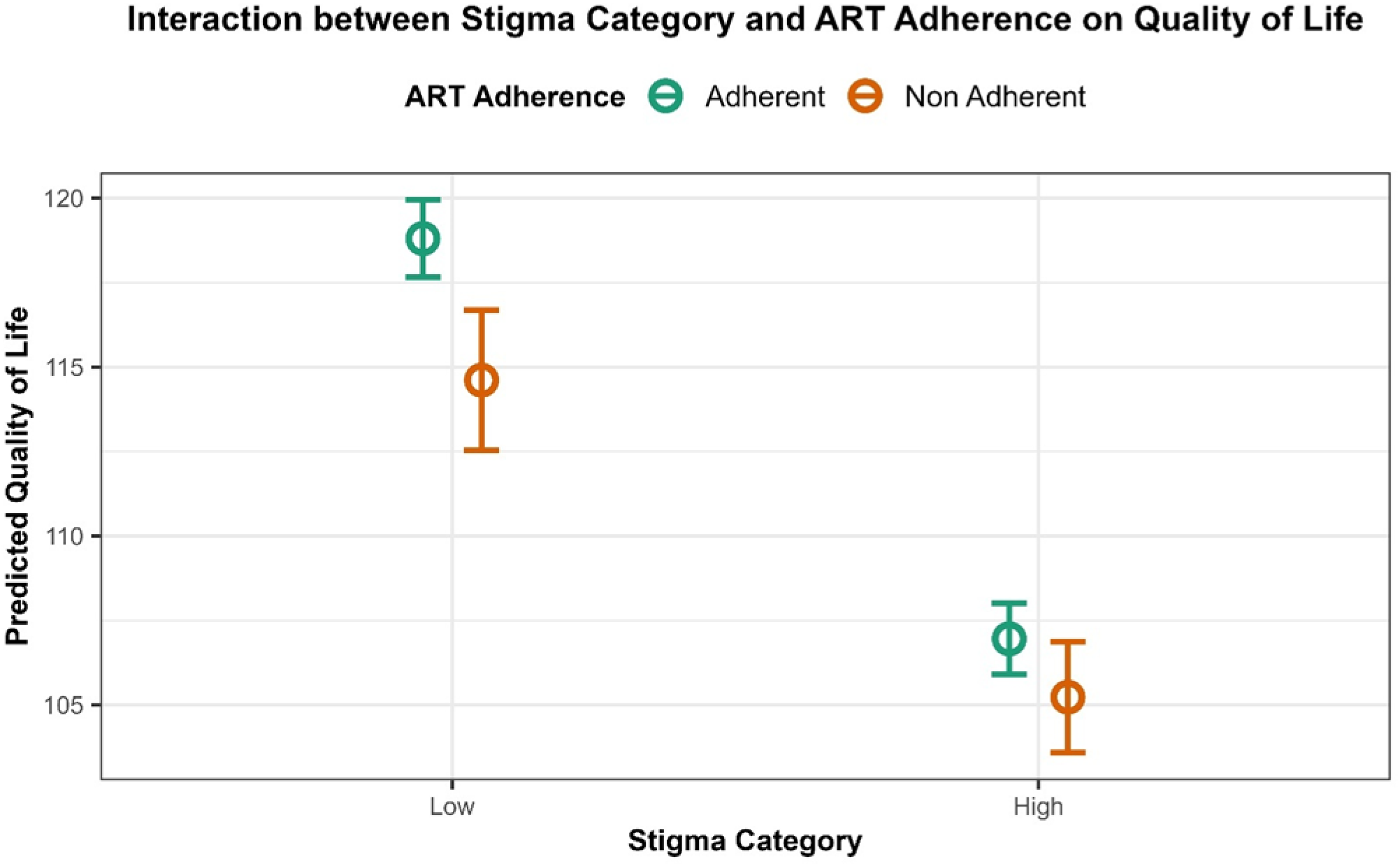
Adjusted predicted total domain-based quality-of-life score by internalized stigma and overall ART adherence among women living with HIV

## Discussion

In this study of WLHIV receiving routine care in Lagos, Nigeria, more than half of participants had high internalized stigma, and women with high stigma had consistently poorer HRQoL across all WHOQOL-HIV BREF domains. In the final multivariable model, internalized stigma remained the strongest independent correlate of lower total domain-based QoL score, while overall ART non-adherence was also independently associated with poorer QoL. Taken together, these findings support the central premise of this study: that wellbeing among WLHIV is shaped not only by clinical management, but also by psychosocial experiences that influence treatment engagement and everyday functioning. This overall pattern is consistent with previous evidence showing that HIV-related self-stigma is negatively associated with quality of life across different settings, including among WLHIV in India and among adults living with HIV in Finland.^25–27^

The most important finding in the present study was the strength and consistency of the association between internalized stigma and poorer QoL. Women with high internalized stigma had lower scores not only for the primary outcome, total domain-based QoL, but also across all six WHOQOL-HIV domains. This suggests that internalized stigma is more than a social label or attitudinal burden; it is a multidimensional psychosocial experience that shapes how WLHIV perceive their physical health, emotional wellbeing, autonomy, social connectedness, environment, and personal meaning.^52,53^ Similar findings have been reported in South India, where stigma among WLHIV was linked to poorer adherence and poorer quality of life, and in Finland, where self-stigma was negatively associated with multiple domains of HRQoL.^25,26^ The consistency of these findings across very different cultural and health-system contexts strengthens the interpretation that internalized stigma is a major determinant of wellbeing among women living with HIV.^15^

The domain-specific pattern observed in this study adds important nuance. The largest descriptive differences were seen in the psychological, environmental, and spirituality/religion/personal beliefs domains, suggesting that internalized stigma may be especially damaging in areas linked to self-acceptance, emotional security, perceived safety, and life meaning. For WLHIV, this is particularly plausible because stigma may intersect with caregiving roles, relationship dynamics, disclosure pressures, and fear of social judgment.^54,55^ A systematic review focused on women living with HIV similarly concluded that stigma has broad and often disproportionate consequences for women’s HRQoL, especially through psychosocial pathways.^15^ In the Nigerian context, this interpretation is also consistent with review evidence showing that stigma remains a major barrier to HIV care engagement, disclosure, and psychosocial wellbeing despite the existence of relevant policies and programmes.^22,56^

ART non-adherence was also independently associated with lower total domain-based QoL score. This finding suggests that adherence should not be viewed only as a biomedical behavior tied to viral suppression, but also as a marker of treatment confidence, daily functioning, and continuity of care. WLHIV who maintain high adherence may experience better health stability, fewer treatment interruptions, and a greater sense of control over their illness, all of which may contribute to better quality of life. Conversely, poor adherence may reflect unresolved psychosocial distress, stigma-related disengagement, competing household demands, or barriers to sustained care. This interpretation is consistent with previous work linking stigma, adherence, and QoL among WLHIV in India,^26^ with evidence that stigma can undermine adherence and retention in care more broadly,^21^ and with emerging intervention evidence showing that stigma-reduction approaches incorporating psychoeducation, counselling, and coping support can also improve ART uptake and adherence.^29^ It is also consistent with evidence showing that quality of life among women living with HIV is shaped by treatment-related functioning and psychosocial stressors in real-world settings.^27,57,58^

The interaction analysis adds another layer to the interpretation of these findings. Predicted total domain-based QoL scores were highest among women with low internalized stigma who were adherent to ART and lowest among those with high stigma who were non-adherent. Across both adherence strata, higher stigma remained associated with poorer predicted QoL, while across both stigma strata, adherence was associated with better predicted QoL. Although this pattern should be interpreted cautiously unless the formal interaction term is statistically significant, it is conceptually plausible. Internalized stigma may reduce QoL directly through shame, fear, and social withdrawal, while also operating indirectly through treatment-related behaviors, coping, and engagement with care. This is in line with prior studies suggesting that stigma can affect wellbeing both directly and through adherence-related pathways,^25,26^ with more recent evidence linking stigma to poorer mental and physical health, decreased social support, and reduced overall quality of life,^59,60^ and with work describing self-stigma as part of the pathway from HIV-related stigma to quality of life.^61,62^

Beyond stigma and adherence, several structural and psychosocial factors were independently associated with total domain-based QoL. Older age was associated with slightly lower QoL, while living more than 5 km from the health facility was associated with poorer QoL and living closer was associated with better QoL. Women in the middle-income category and those who preferred not to disclose their income also had lower QoL scores than the reference group. These findings suggest that HIV-related wellbeing is shaped not only by psychosocial burden, but also by the practical burden of navigating care. Distance to the facility may reflect transport difficulties, time costs, and reduced convenience in sustained care engagement, while income-related patterns may reflect financial vulnerability or reluctance to disclose sensitive socioeconomic information. This interpretation is consistent with broader evidence that financial constraints, transportation barriers, and service accessibility can influence HIV-related stress, help-seeking, and overall wellbeing.^63–65^ For example, qualitative work from Cameroon showed that economic vulnerability and access barriers affected whether people with HIV could obtain needed support within care systems.^66^ A broader review of determinants of HRQoL among people living with HIV also found that sociodemographic and socioeconomic factors, including income and access-related constraints, are important correlates of wellbeing.^48^

Social support was positively associated with total domain-based QoL in the adjusted model. This finding aligns with wider evidence that supportive relationships can buffer the effects of stigma and promote better functioning among people living with HIV. In Ghana, social support is positively associated with QoL, reinforcing the role of relational resources as protective factors in HIV care.^28^ This is also consistent with broader evidence linking stigma, social support, and quality of life among women and other people living with HIV.^15,67^ In the present study, this is especially important because WLHIV often navigate layered pressures related to disclosure, caregiving, partnership dynamics, and community expectations. Strengthening support through peer groups, family-based approaches, or clinic-linked psychosocial services may therefore represent a practical pathway for improving wellbeing alongside biomedical care.

One finding that warrants cautious interpretation is the positive association between recent opportunistic infection history and higher total domain-based QoL score. This direction is counterintuitive and should not be interpreted as evidence of benefit. Because the present analysis is cross-sectional and based on baseline data, this association may reflect residual confounding, timing of illness and recovery, survivorship effects, or greater healthcare contact among women who had recently experienced illness and were subsequently receiving closer monitoring or support. Existing literature more commonly reports the opposite pattern, with opportunistic infections generally associated with poorer HRQoL among people living with HIV, suggesting that the present result is unlikely to reflect a true beneficial effect.^68,69^ Rather than viewing this as contradictory to existing knowledge, it is more appropriate to treat it as hypothesis-generating and deserving of further evaluation in longitudinal analyses.

These findings have important implications for HIV care delivery. They suggest that improving QoL among WLHIV requires more than sustained ART provision alone. Internalized stigma appears to be a central and potentially modifiable driver of poorer wellbeing, while adherence and social support remain important linked mechanisms. For practice, this supports integrating stigma screening, psychosocial counselling, adherence support, and women-centred supportive interventions into routine HIV care. This is particularly relevant in Nigeria, where review evidence indicates that the challenge is not only the existence of policy, but the implementation of context-specific stigma-reduction strategies within real-world care systems.^22^ Recent evidence on stigma-reduction interventions also suggests that combined behavioural and psychosocial approaches may be especially useful when the goal is to improve both treatment outcomes and lived experience.^29,70,71^ This broader direction is also consistent with evidence and guidance emphasizing the importance of integrating psychosocial and mental health support into HIV service delivery and of developing supportive interventions specifically responsive to the needs of women living with HIV.^58,72,73^

This study has several strengths. It used validated measures of internalized stigma and HIV-related QoL, assessed adherence using a composite measure derived from pharmacy refill and self-report, and examined QoL as a multidimensional outcome rather than relying solely on a single global self-rating.^17,23,24,42^ The relatively large sample also improved the precision of estimates. At the same time, several limitations should be acknowledged. Because this was a baseline cross-sectional analysis, causal inferences cannot be made about the direction of associations, and the use of baseline observational data nested within a quasi-experimental study should be interpreted as descriptive of relationships prior to intervention exposure rather than as evidence of causality.^74,75^ The study was conducted at a single HIV treatment centre, which may limit generalizability to other settings. Some measures, particularly self-reported adherence and psychosocial variables, may also have been influenced by reporting bias, a limitation long recognized in self-reported adherence measurement.^47^ In addition, dichotomising internalised stigma may have reduced the nuance in the distribution of exposure. Nevertheless, the consistency of the observed associations across multiple quality-of-life domains strengthens confidence in the overall interpretation.

## Conclusion

Internalized stigma and ART non-adherence were independently associated with poorer total domain-based QoL among WLHIV in Lagos, Nigeria. Internalized stigma emerged as the strongest correlate of poorer wellbeing, underscoring the need to move beyond narrowly biomedical models of HIV care. Interventions that combine stigma reduction, adherence support, and psychosocial strengthening may be particularly valuable for improving patient-centered outcomes among WLHIV in low-resource settings.

## Data Availability

De-identified data are not publicly available due to participant confidentiality. Access may be requested from the Nigerian Institute of Medical Research Institutional Review Board, subject to ethical approval.

## Supplementary Files

Supplementary File 1. Baseline questionnaire and data collection instrument

Supplementary File 2. Variable definitions, scoring procedures, and model-building strategy

Supplementary Table S1. Expanded baseline characteristics of women living with HIV by internalized stigma category

Supplementary Table S2. Unadjusted and intermediate multivariable linear regression models for factors associated with total domain-based quality-of-life score

Supplementary Table S3. Interaction model for internalized stigma and overall ART adherence in relation to total domain-based quality-of-life score

## Supplementary Appendix 1. Variable definitions, scoring, and model-building strategy

### Internalized HIV stigma

Internalized stigma was assessed using the 10-item Internalized Stigma of HIV/AIDS Tool (ISAT). Each item measured negative self-perceptions related to living with HIV, including shame, guilt, worthlessness, concealment, inferiority, self-blame, and perceived burden. In the analytical dataset, responses were scored on a 5-point Likert scale, with higher scores indicating greater internalized stigma. Item scores were summed to generate a total internalized stigma score. For descriptive and regression analyses, the total score was dichotomized at the sample median into low internalized stigma (<20) and high internalized stigma (≥20). Internal consistency of the scale in the study sample was excellent (Cronbach’s alpha = 0.95).

### ART adherence

ART adherence was assessed using two complementary approaches: pharmacy refill adherence and self-reported adherence.

### Pharmacy refill adherence

Pharmacy refill adherence was derived from pharmacy refill records and represented the proportion of prescribed ART medication obtained within the expected refill interval. A continuous adherence score was first computed and then converted to a percentage. For descriptive analyses, refill adherence was categorized as:

- Good adherence: score ≥2.5 (approximately ≥83%)
- Fair adherence: score 1.5–2.0 (approximately 50%–67%)
- Poor adherence: score <1.5 (<50%)

For binary analyses, pharmacy refill adherence was dichotomized as:

- Adherent: ≥95%
- Non-adherent: <95%

Self-reported adherence

Self-reported adherence was assessed using questions on missed doses, medication-taking consistency, and dose timing. Responses were scored on a 0–4 scale. For descriptive analyses, scores were categorized as:

- Adherent: score ≥3.5
- Non-adherent: score <3.5

For the primary binary adherence variable, participants were classified as adherent if they reported all of the following:

1. never missing a dose, or missing no more than one dose in the previous month,
2. taking all or most of their medication as prescribed, and
3. taking doses on time or within one hour of the prescribed time. Participants who failed to meet all criteria were classified as non-adherent.

### Composite overall ART adherence

A composite overall adherence variable was created to improve ascertainment of treatment-taking behavior. Participants were classified as adherent only if they met the ≥95% adherence threshold on both pharmacy refill adherence and self-reported adherence. Participants who did not meet the threshold on either or both measures were classified as non-adherent. The adherence section demonstrated acceptable internal consistency (Cronbach’s alpha = 0.77).

### Quality of life

Health-related quality of life was measured using the WHOQOL-HIV BREF. The instrument assessed six domains:

1. physical health,
2. psychological health,
3. level of independence,
4. social relationships,
5. environment, and
6. spirituality/religion/personal beliefs.

Negatively phrased items were reverse-coded before scoring. Domain scores were calculated by averaging the relevant item scores within each domain and multiplying by four, yielding domain scores ranging from 4 to 20, with higher scores indicating better quality of life.

For this study, the primary outcome was the total domain-based quality-of-life score, calculated as the sum of the six WHOQOL-HIV BREF domain scores. The two general items on overall quality of life and overall health were summarized descriptively but were not used as the primary outcome in regression analyses.

### Covariates

Candidate covariates were selected based on conceptual relevance and availability in the baseline dataset. These included age, marital status, education level, monthly income, religion, number of children, living arrangement, residential area, distance to the health facility, health insurance status, disclosure status, social support, time since HIV diagnosis, current ART use, viral load category, most recent viral load, WHO clinical stage, opportunistic infection history, and BMI category.

### Model-building strategy

The main inferential analysis assessed the association of internalized stigma and overall ART adherence with total domain-based QoL score using linear regression. A sequential modelling strategy was applied.

In Model 1, each candidate predictor was entered separately to estimate unadjusted associations with total domain-based QoL score. Variables associated with the outcome at p ≤ 0.05 in unadjusted analyses were entered simultaneously into Model 2, which estimated adjusted associations. Variables that remained statistically significant at p ≤ 0.05 in Model 2 were retained in Model 3, which served as the final parsimonious model. Effect estimates were reported as beta coefficients (β) with 95% confidence intervals (CIs).

### Interaction analysis

To assess whether the association between internalized stigma and total domain-based QoL score differed by ART adherence status, a stigma-by-adherence interaction term was included in a multivariable linear regression model. The interaction model was specified as:

QoL = β₀ + β₁(stigma category) + β₂(ART adherence) + β₃(stigma category × ART adherence) + ε where β₃ represents the interaction effect between internalized stigma and overall ART adherence on total domain-based QoL score. Adjusted predicted means and 95% confidence intervals were estimated for each stigma–adherence combination and displayed graphically.

**Supplementary Table S1.** Expanded baseline characteristics of women living with HIV by internalized stigma category.

| Characteristic | High internalized stigma (n = 564) | Low internalized stigma (n = 441) | Overall (N = 1,005) | p-value |
| --- | --- | --- | --- | --- |
| Religion |  |  |  | 0.215 |
| Christian | 484 (85.8) | 391 (88.7) | 875 (87.1) |  |
| Islam | 80 (14.2) | 50 (11.3) | 130 (12.9) |  |
| Number of children, mean (SD) | 2.3 (1.6) | 2.3 (1.8) | 2.3 (1.7) | 0.998 |
| Living arrangement |  |  |  | 0.528 |
| Living alone | 61 (10.8) | 45 (10.2) | 106 (10.5) |  |
| Living with parents | 26 (4.6) | 22 (5.0) | 48 (4.8) |  |
| Living with children only | 88 (15.6) | 52 (11.8) | 140 (13.9) |  |
| Living with extended family/friends | 36 (6.4) | 30 (6.8) | 66 (6.6) |  |
| Living with spouse/partner | 352 (62.4) | 292 (66.2) | 644 (64.1) |  |
| Health insurance status |  |  |  | 0.376 |
| No – pay out of pocket | 540 (95.7) | 416 (94.3) | 956 (95.1) |  |
| Yes – enrolled in NHIS or other scheme | 24 (4.3) | 25 (5.7) | 49 (4.9) |  |
| WHO clinical stage, mean (SD) | 1.0 (0.2) | 1.0 (0.1) | 1.0 (0.2) | 0.456 |
| Most recent viral load, mean (SD) | 23,764.9 (425,480.4) | 5,465.2 (64,514.1) | 15,734.9 (321,593.5) | 0.314 |
| Pharmacy refill adherence |  |  |  | 0.098 |
| Adherent | 504 (89.4) | 378 (85.7) | 882 (87.8) |  |
| Non-adherent | 60 (10.6) | 63 (14.3) | 123 (12.2) |  |
| Self-reported adherence |  |  |  | <0.001 |
| Adherent | 392 (69.5) | 350 (79.4) | 742 (73.8) |  |
| Non-adherent | 172 (30.5) | 91 (20.6) | 263 (26.2) |  |

**Supplementary Table S2.** Unadjusted and intermediate multivariable linear regression models for factors associated with total domain-based quality-of-life score among women living with HIV.

| Predictor | Category / comparison | Model 1: Unadjusted $\beta$ (95% CI) | p-value | Model 2: Adjusted $\beta$ (95% CI) | p-value |
| --- | --- | --- | --- | --- | --- |
| Age | Per 1-year increase | -0.064 (-0.124 to -0.004) | 0.038 | -0.08 (-0.14 to -0.01) | 0.017 |
| BMI category | Underweight (Ref) |  |  |  |  |
|  | Normal weight vs underweight | 0.560 (-0.940 to 2.059) | 0.464 | 3.68 (1.34 to 6.03) | 0.002 |
|  | Overweight vs underweight | 3.597 (0.868 to 6.326) | 0.010 | -2.17 (-4.04 to -0.30) | 0.023 |
|  | Obese vs underweight | -1.992 (-4.194 to 0.210) | 0.076 | 1.23 (-0.04 to 2.50) | 0.058 |
| Monthly income | ₦10,000–₦49,999 (Ref) |  |  |  |  |
|  | Less than ₦10,000 vs Ref | 1.250 (-1.906 to 4.405) | 0.437 | -2.18 (-5.04 to 0.68) | 0.135 |
|  | ₦50,000–₦99,999 vs Ref | -2.041 (-4.480 to 0.399) | 0.101 | -2.48 (-4.58 to -0.38) | 0.021 |
|  | ₦100,000 and above vs Ref | 1.514 (-0.995 to 4.023) | 0.237 | 1.16 (-1.03 to 3.35) | 0.299 |
|  | Prefer not to say vs Ref | -3.675 (-5.968 to -1.382) | 0.002 | -2.17 (-4.21 to -0.14) | 0.037 |
| Distance to health facility | 1–5 km (Ref) |  |  |  |  |
|  | <1 km vs 1–5 km | 2.907 (1.197 to 4.617) | <0.001 | 1.96 (0.45 to 3.47) | 0.011 |
|  | >5 km vs 1–5 km | -6.122 (-8.104 to -4.139) | <0.001 | -4.28 (-6.14 to -2.41) | <0.001 |
| Residential area | Rural (Ref) |  |  |  |  |
|  | Semi-urban vs rural | 7.908 (4.645 to 11.171) | <0.001 | 1.32 (-1.59 to 4.23) | 0.373 |
|  | Urban vs rural | 8.542 (5.572 to 11.511) | <0.001 | 1.63 (-1.11 to 4.37) | 0.242 |
| Number of children | Per child increase | -0.469 (-0.910 to -0.029) | 0.037 | -0.24 (-0.69 to 0.21) | 0.299 |
| Opportunistic infection history | Yes vs No | 8.805 (5.716 to 11.894) | <0.001 | 4.90 (2.11 to 7.68) | <0.001 |
| Social support | Any support vs no support | 4.771 (2.932 to 6.611) | <0.001 | 2.23 (0.58 to 3.88) | 0.008 |
| Overall ART adherence | Non-adherent vs adherent | -3.554 (-5.246 to -1.861) | <0.001 | -2.76 (-4.23 to -1.29) | <0.001 |
| Internalized stigma | High vs low | -11.371 (-12.719 to -10.023) | <0.001 | -9.96 (-11.31 to -8.61) | <0.001 |

**Supplementary Table S3.**
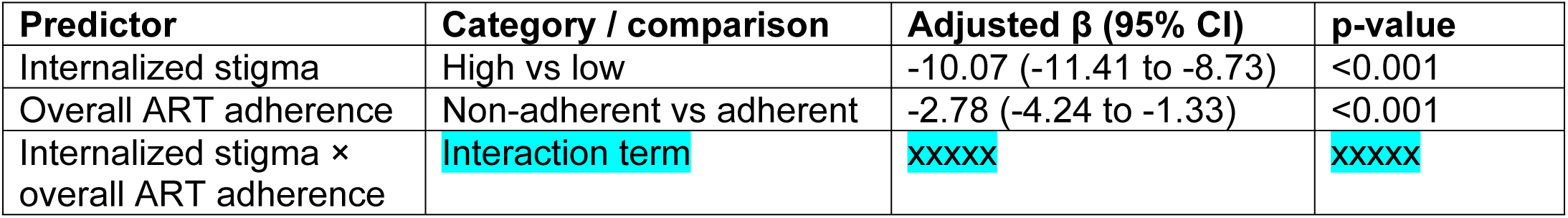
Interaction model for internalized stigma and overall ART adherence in relation to total domain-based quality-of-life score among women living with HIV.

